# Oxygen needs in austere environment : French military health service experience

**DOI:** 10.1101/2020.04.06.20055780

**Authors:** Pierre-Julien Cungi, Quentin Mathais, Erwan D’Aranda, Mickael Cardinale, Philippe Goutorbe, Julien Bordes, Eric Meaudre

## Abstract

**INTRODUCTION:** Management of Oxygen supplies is a recurring problem for the intensivists and anesthesiologists working in an austere environment. The French military health service has chosen oxygen concentrators (OC) as the primary source of oxygen.

**OBJECTIVES:** The main objective was to evaluate the feasibility of using OC as the main source of 0_2_ for intensive care patients. We assess the need to use pressurized 0_2_ during the ICU hospitalization. The secondary objectives were to identify the causes of the use of pressurized 0_2_.

**MATERIAL AND METHOD:** We realize an interventional cohort study at the French role 3 hospitals located in the Republic of Djibouti. The criteria of inclusion were all patients aged over 18 years, requiring oxygen and admitted to intensive care.

**RESULTS:** We include 35 patients over 6-month period for 251 days of oxygenation, including 142 days of invasive mechanical ventilation. The population include 21 (60%) men, aged of 35 (30 - 49) years. Twenty-eight (80%) patients benefits of invasive ventilation. Median 0_2_ administration duration was 6 (3-10) days, and the median duration of mechanical ventilation was 3 (1-5) days. Nineteen pressurized O2 treatments were required over 251 days of oxygen therapy, or 7.5% of the total oxygen therapy time. The causes of recourse were in 10 cases (52.6%) severe ARDS, in 6 cases (31.6%) an emergency orotracheal intubation and in 3 cases (15.8%) a transfer. Only one OC dysfunction occurred during the study.

**CONCLUSION:** OC can be used as a primary source for intensive care patients in an austere environment. The use of pressurized 0_2_ remains imperative in the event of an electrical failure and the need to use high Fi0_2_ over 60%.

## Introduction

Oxygen delivery for the ventilated patient can be challenging in an austere environment, in developing countries or during disaster situation. Standard medical oxygen (99.9% 0_2_) is available in spressurised cylinders or hospital wall-mounted distribution system. This spressurised oxygen has the advantage of providing a high flow of 0_2_ and a high concentration, which are essential for the functioning of intensive care ventilators and allows doing high flow oxygen therapy. For logistic and cost reasons, oxygen cylinders and pressurized in-hospital oxygen delivery systems are not always available in austere environment.

One solution to providing medical oxygen (93% 0_2_) is to use an oxygen concentrator (OC), which needs a low electrical power to generate high concentrations of oxygen from the air at flow rates from 0.5 liters per minute (lpm) to 10 lpm. These flow rates are sufficient for the use of a nasal cannula or facial mask in a patient with moderate lung function impairment.

The WHO recommends the use of OC as a primary source of oxygen in the developing country because of its low cost (5$/m3 versus 30$/m3 for spressurised 0_2_) and the limited logistic needs(1). The use of OC is already described for standard medical care and in anesthesia(2,3). There is no documentation about the use of OC in the intensive care unit for patients requiring mechanical ventilation. This usage is more complex and needs to use a turbine ventilator with a low-pressure oxygen inlet and not a ventilator using spressurised gas as the motor fluid(4,5). Also, the Fi0_2_ delivered to patients depends on the minute volume and bypass flow of the ventilator.

French Army is often engaged in peacekeeping and support missions in Africa (“Operation Barkhane”) and several Forward Surgical Unit (FSU) are deployed to support French and allied forces. A FSU is a light mobile structure consisting of two tents with their electrical power supplies which is deployed in austere environment and must be autonomous for weeks. It allows to perform damage control surgery and intensive care closer to the combat field with a small team (11 peoples). These units can do 20 surgeries a day during 4 days and maintain patients in intensive care for 48h waiting for evacuation to a French role 4 hospitals. FSU dispose of 21,6 m3 of pressurised 0_2_ in cylinders (10 cylinders of 3l and 6 cylinders of 6l pressurize at 200 bar). This amount of oxygen is sufficient to maintain only two patients with FiO2 = 0.5 under mechanical ventilation for 48h. Lung function impairment is a frequent problem for the FSU. Since 2012, 57 patients (32 war injury, 17 severe trauma, and 7 medical pathologies) were evacuated from war theater with ARDS criteria(6). Theses patient often need a 48 h field critical care in the FSU before been transferred in a role 3 or role 4 structure.

We have designed a feasibility study to describe the usage of the couple ventilator + OC and determine the limits of the use of extractors as the primary source of oxygen in the intensive care unit of our role 3 located in Djibouti.

## Methods

We designed an observational prospective monocentric study in the role 3 “Surgical and Medical hospital Bouffard” in Djibouti (Djibouti). This hospital serves as a support facility for the French forces deployed in Djibouti and the Middle East, and it includes a medical and surgical service with 20 hospital beds, 8 intensive care beds, a theater room with 2 operating rooms, and of emergency service. In the GMC, the ruse il to use the OC as the primary source of oxygen in intensive care, anaesthesia, emergency room, and wards.

*The primary endpoint* was to evaluate the need for pressurized oxygen for patients who were treated by oxygen on OC.

*The secondary endpoints* were the identification of the causes of the use of pressurized O_2_, the description of the pulmonary function during the use of oxygen therapy, an estimation of the amount of O_2_ saved, and an evaluation of costs.

### Inclusion and exclusion criteria

The inclusion criteria were all patients over the age of 18 years old requiring oxygen admitted to intensive care on role 3 Bouffard.

### Security criteria

The criteria for the systematic use of pressurised O_2_ were:

- Severe hypoxemia (PaO_2_ / FiO_2_ ratio <100) requiring more oxygen than 10 l/min on the OC despite ventilatory optimization: protective ventilation with recruitment maneuver and titration of the PEEP, use of neuromuscular blocking agents, and prone positioning.
- Situations requiring a high FiO_2_ : pre-oxygenation before intubation, cardio- respiratory arrest or all other situations requiring a flow of O_2_ greater than 10 l / min on the EO.
- OC failure.

### Materials

FiO_2_ was measured using the Datex D-FEND module of the General Electric Datex Omheda monitoring system. The Pulmonetic LTV1000 turbine ventilator was used for this study. OC were SeQual Integra 10-OM (SeQual, San Diego, CA). The LTV 1000 ventilator and Sequal Integra 10-OM OC were selected because the FMHS (French military health service) uses them in FSUs and role 3. The SeQual Integra 10-OM OC is a device capable of generating from 0.5 to 10 L/min of continuous oxygen flow. It is an electronically operated OC that separates oxygen from room air. An indicator light is activated when oxygen concentration falls below preset levels of 85%.

### Data collection

We collected the use of pressurised oxygen and its duration, the causes of use, and the settings of the ventilator at each change. The clinical, biological, and bacteriological data were collected daily by the intensivist. Changes in the chest x-ray or CT scan were collected. The diagnosis of ARDS and its categorisation were made according to the “Berlin definition” after optimisation of the ventilator settings, including PEEP level titration, use of neuromuscular blocking agents, and prone positioning

### Ethics

The study protocol was reviewed and approved by the ethics committee of the Bouffard hospital (Role 3). Written consent from patients was not required as care was not changed.

### Statistics

Continuous numerical variables are expressed in median and interquartile. Qualitative value are expressed in number and percentage. We used Khi2 and exact fisher test for the qualitative value. We used Wilcoxon-Mann-Whitney test for the continuous numerical variable. Statistics were realised with R 3.3.30 (R Foundation for statistical computing). Type I risk was fixed at 5%.

## Results

132 patients were admitted to intensive care over 6 months (between July 2014 and October 2014 then between June 2015 and August 2015), 61 did not require O2, 36 were under 18 years old, and 35 patients were included (Figure 1).

**Figure 1.**
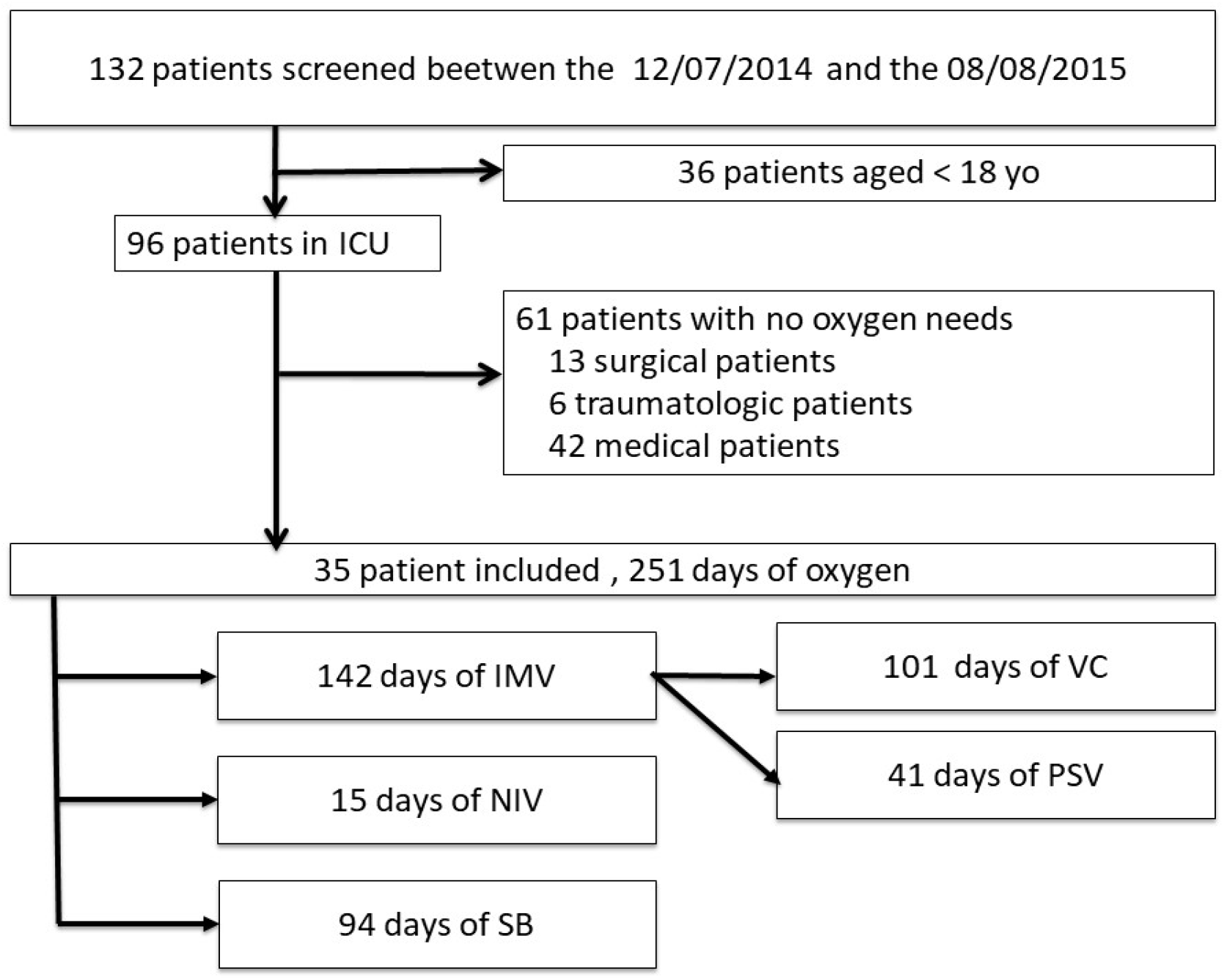
Flow chart. ICU : intensive care unit ; IMV : invasive mechanical ventilation ; VC : volume-controlled ventilation ; PSV : pressure support ventilation ; NIV : non invasive ventilation ; SB : spontaneous breathing.

### Population

The population was composed of 21 (60%) men, aged of 35 (30 - 49) years. The reason for admission was medical in 21 cases (60%), following a trauma in 8 cases (22.9%), and surgical in 6 cases (17.1%). Eight (22.9%) patients died. On admission, the IGS II score was 37 (23 - 51), and the SOFA score was 5 (2 - 10). Detailed characteristics of the patients are shown in Table 1. Twenty-eight (80%) patients were intubated during the ICU stay, and 17 were intubated before admission to the intensive care unit. The intensive care length of stay under oxygen therapy was 6 (3-10) days, and the duration of mechanical ventilation was 3 (1-5) days. Eight (22.8%) patients benefited from NIV including 6 (17%) after extubation. The total number of oxygen days for the 35 patients was 251, including 142 days of invasive ventilation (Volume-Controlled Ventilation and Pressure Support Ventilation), 15 days of NIV, and 94 days of oxygen therapy with a simple mask or nasal cannula.

**Table 1.** Patients characteristics

|  |  |  |
| --- | --- | --- |
| Age | 39 | (15-70) |
| Sex |  |  |
| *Women, n %* | 14 | 40% |
| *Men, n %* | 21 | 60% |
| Weight (kg) | 71 | (35-120) |
| Height (m) | 1,7 | (1,5-1,85) |
| BMI (kg/m<sup>2</sup>) | 25 | (15,5-39) |
| Reason for admission |  |  |
| *Medical* | 6 | 17 % |
| *Surgical* | 21 | 60 % |
| *Traumatology* | 8 | 23 % |
| Duration of O<sub>2</sub> therapy (j) | 6 | (0-35) |
| Death (n - %) | 8 | 22,85% |
|  | 3 |  |
| APACHE II | 19,62 | (2-50) |
| IGS II | 38,8125 | (8-80) |
| SOFA | 5 | (2-10) |
| Blood Gas |  |  |
| pH | 7,27 | (6,8 - 7,48) |
| PaO<sub>2</sub> (mmHg) | 100 | (46 - 200) |
| PaCO<sub>2</sub> (mmHg) | 36 | (13 - 101) |
| HCO<sub>3</sub><sup>-</sup> (mmHg) | 19 | (0 - 39) |
| O<sub>2</sub> flow (l/min) | 2,5 | (0 - 15) |

BMI : Body mass index ; APACHE II : Acute Physiology And Chronic Health Evaluation II ; IGS II ; IGS II : Simplified gravity index II ; SOFA : Sequential Organ Failure Assessment.

### Primary endpoint

We had to use pressurised O_2_ 19 days (7.5%) over the 251 days of oxygen therapy. The use of pressurised O_2_ only concerned intubated patients in volume controlled mechanical ventilation(19% of mechanical ventilation days).

### Cause and duration of use of pressurised O_2_

The causes of use of pressurised O2 are an ARDS for 10 (53%) uses, a preoxygenation for emergency intubation for 5 (26%) uses, a transport for 3 (16 %) use and one failure of OC (5%). The duration of pressurised oxygen use was less than 4 hours in 9 cases (47.3%): IOT in emergency for 5 cases (55.6%), transfer to radiology or theater room in 3 cases (33.3%) and for an ARDS in 1 case (11.1%). In 10 cases (52.6%) the duration was more than 4 hours (long-term remedy): ARDS for 9 cases (90%) and a failure of the OC in 1 case (10%). ARDS criteria were more frequent on the days of use of pressurised O_2_ compared to days with OC oxygen therapy: (52.6%) in the group O_2_ pressurised versus 3 % in the OC group (p <0.001). The different parameters are described in Table 2.

**Table 2.** Characteristics of ventilation for patients wich request spressurised oxygen use.

|  | >4h | < 4h |  |
| --- | --- | --- | --- |
| Pressurized O2 request (n) | 10 | 9 |  |
| FiO<sub>2</sub> (%) | 84 | 47 | p<0,005 |
| PaO<sub>2</sub> (mmHg) | 128 | 119 | NS |
| PaCO<sub>2</sub> (mmHg) | 52 | 40 | NS |
| Peep (cmH<sub>2</sub>O) | 7 | 8 | NS |
| MV (l/min) | 8,1 | 7,5 | NS |
| PaO<sub>2</sub> /fiO<sub>2</sub> | 123 | 248 | p<0,001 |
| ARDS | 8 | 1 | p<0,001 |
| SOFA | 12 | 8 | NS |

Peep : positive end expiratory pressure ; MV : minute ventilation ; ARDS : Acute respiratory distress syndrome; SOFA : Sequential Organ Failure Assessment.

### Description of the pulmonary function of patients

Patient which required pressurised O_2_ had a significantly altered pulmonary function with oxygenation trouble: Pa02/Fi0_2_ = 185 on Pressurized 0_2_ group versus 385 on Oxygen concentrator group (p<0.001) for a respective Fi02 of 68% and 38%. The lung compliance was reduced (34 mL/CmH_2_0 vs. 47 mL/cmH_2_0 – p < 0.001) and the minute volume need for a correct decarboxylation were high (9,2 l/min vs. 7,6 l/min – p < 0.05). The detail of the pulmonary function is shown in table 3.

**Table 3.** Pulmonary function during days of pressurised oxygen use on the patient under mechanical ventilation.

|  | OC days | O<sub>2</sub> Pressurised days |  |
| --- | --- | --- | --- |
| Oxygenation days (d) | 82 | 19 |  |
| Days with ARDS criterion (d) | 4 | 10 | < 0,001 |
| FiO<sub>2</sub> (%) | 38 | 68 | < 0,001 |
| Pplat (cmH<sub>2</sub>O) | 17,5 | 23,5 | < 0,001 |
| Ppeak (cmH<sub>2</sub>O) | 25 | 36 | < 0,001 |
| Peep (cmH<sub>2</sub>O) | 6 | 7,6 | < 0,05 |
| MV (l/min) | 7,9 | 7,6 | NS |
| MV cor (l/min/mmHg) | 7,6 | 9,2 | < 0,05 |
| Comp (ml/cmH<sub>2</sub>O) | 47 | 34 | < 0,01 |
| pH | 7,37 | 7,26 | < 0,01 |
| PaO<sub>2</sub> (mmHg) | 131,8 | 117,5 | < 0,05 |
| PaCO<sub>2</sub> (mmHg) | 39,5 | 47,5 | < 0,05 |
| PaO<sub>2</sub> /FiO<sub>2</sub> | 348 | 185 | < 0,001 |
| Comp : thoracic compliance ; Pplat : plateau pressure ; Ppeak : Peak pressure ; Peep : end expiratory positive pressure ; MV : Minute Volume ;MV cor : MV normalised for a 40 mmHg PaCO<sub>2</sub>. |  |  |  |

### Economic consideration

the total amount of oxygen produced by the OC during this study is estimated at 1040 m3 of O_2_. Based on WHO data, the cost of cylinder oxygen ranges from US $ 10 to US $ 30 per m3 (without the transportation of cylinders) versus US $ 2 to US $ 8 per m3 for an EO(1). This cost estimate assesses the savings made between $ 10000 and $ 30000 for the six months of the study.

### Reliability of concentrator

we noticed one technical failure of an OC (FiO_2_ alarm produced weak) was detected when it was started and led to leaving the patient under pressurised O_2_.

## Discussion

This study is the first using OC in real condition in austere environment. We showed that OC as primary source of oxygen in ICU is safe for oxygenation of ICU patients for the three main modalities of administration: Facial mask, non-invasive ventilation and invasive mechanical ventilation. The previous description of use in ICU was reports cases, and the limits of the couple OC – turbine ventilator studies were studied on benchmark for some models of ventilator (4,5,7–10).

The pressurised source of O2 concerned specific and brief situations for halves of them like transport or pre-oxygenation before intubation. The prolonged use of pressurised 0_2_ mainly concerned patients with severe lung function impairment (severe ARDS of the Berlin classification). Only one OC dysfunction occurred during the study despite the austere climatic conditions of the Horn of Africa (local temperature greater than 30 ° C and hygrometry level greater than 80% in the intensive care ward).

The WHO recommends OC as the primary source of O_2_ in developing countries in “Technical specification for oxygen concentrator, published in 2015(1). This guide discusses the practice of anesthesia and neonatal CPAP using an OC, but not intensive care for which no data was available until now.

The OC provides normobaric 02, and we can not use the pressure of the fluid to create driving pressure in critical care ventilators, so the use of OC for mechanical ventilation requires the use of turbines ventilators. The consequence of this association is the variation in FiO_2_ as a function of the patient’s minute ventilation. Previous studies have shown that the FiO_2_ delivered depends mainly on the oxygen flow rate of the OC, the minute volume, the Inspiratory (time)/ Expiratory time ratio, the bypass flow, and that it was necessary to monitor the delivered FiO_2_ on the inspiratory branch of the ventilator. In the absence of a measuring cell, it is possible to estimate the Fi02 administered to the patient using the charts produced by *Bordes et al* from the O2 flow rate of OC and the minute volume(4). The Fi02 delivered for a same normobaric 0_2_ flow depends on the model of ventilator due to the design of the gas blender and bypass flow (for example 10 l/min for a pulmonetic LTV1000 and 5 l/min for a ResMed Elisée 350). The user should be aware of these problems and need to train with the couple turbine ventilator / OC and define the possibilities and limits of their equipment to limit the risks for the patients.

The pitfalls of critical care in an austere environment are few explored in the medical literature. Intensive care in developing countries is limited by the varying access to resources from one country to another and a very different financial capability. In developing countries, intensive care beds represent only 2-3% of total hospital beds, compared to more than 10% in industrialised countries(11). The critical care equipment, the training and skills of personnel, the maintenance of equipment, and the intensive care services of developing countries, have a very heterogeneous level.

OC can be a good source of O_2_ in countries with limited resources, because they only require a low-power source of electricity to operate and that their initial purchase costs are relatively low ($ 250 to $ 2,000)(1,12–14). However, these costs are to be compared with the budget allocated to the health of African states. Buying a single OC can represent a significant budget. For example, the budgets for France in 2013 were $ 4,864 /inhabitant versus $ 137 /i in Djibouti, $ 87 / i in Côte d’Ivoire, $ 53 / i in Mali, $ 27 / i in Niger, $ 37 /i in Chad, $ 13 / i in the Central African Republic (source : https://web-prod.who.int/health-financing/repository-of-health-budgets).

The oxygen stocks deployed with an FSU are limited 21.6 m3 of normobaric oxygen in pressurised cylinders. These stocks allow ventilating a patient with a FiO2 at 100% or two patients with a FiO2 at 50% for 48h. These stocks are insufficient to allow the treatment of massive casualties of wounded and maintain several patients in critical care for 48h waiting for extraction. The use of OC coupled with turbine ventilator could be a simple solution for deploying ICU bed with ventilation capacity in austere condition, with a minimum of logistic impact.

These results are even more interesting in the context of the COVID-19 pandemic. Our results suggest that we can use OC associated with turbine ventilator to create ventilated beds without a source of wall pressurised oxygen or a heavy logistic to supply oxygen bottles. However, OC are not sufficient for the patient with a severe alteration of pulmonary function wich need high FIO_2_ (above 60-70%).

## Conclusion

This study confirms the possibility of using OC as the primary source of oxygen in intensive care and pressurised oxygen can be used only for precise needs. The OC can produce 02 with only a low power electrical generator and don’t need a complex logistic circuit.

In mechanical ventilation, the use of EO requires the use of turbine ventilator that does not require pressurised O_2_ to generate a tidal volume.

Pressurised O2 remains essential in 3 clinical situations: situations requiring a FiO_2_ higher than 60-70%, including severe ARDS, emergency intubations, and the transport of ventilated patients.

## Data Availability

Data are available if requested.

